# Transcriptomic meta-analysis of non-Hodgkin’s B-cell lymphomas reveals reliance on pathways associated with the extracellular matrix

**DOI:** 10.1101/2022.04.28.22274444

**Authors:** Naomi Rapier-Sharman, Jeffrey Clancy, Brett E. Pickett

**Author notes:** Corresponding author, address: 3141 LSB Provo, UT 84602-1050.

## Abstract

Approximately 450,000 cases of Non-Hodgkin’s lymphoma are diagnosed annually worldwide, resulting in ∼240,000 deaths. An augmented understanding of the common mechanisms of pathology among relatively large numbers of B-cell Non-Hodgkin’s Lymphoma (BCNHL) patients is sorely needed. We consequently performed a large transcriptomic meta-analysis of available BCNHL RNA-sequencing data from GEO, consisting of 322 relevant samples across ten distinct public studies, to find common underlying mechanisms across BCNHL subtypes. The study was limited to GEO’s publicly available human B-cell RNA-sequencing datasets that met our criteria, and limitations may include lack of diversity in ethnicities and age groups. We found ∼10,400 significant differentially expressed genes (FDR-adjusted p-value < 0.05) and 33 significantly modulated pathways (Bonferroni-adjusted p-value < 0.05) when comparing lymphoma samples to non-diseased samples. Our findings include a significant class of proteoglycans not previously associated with lymphomas as well as significant modulation of extracellular matrix-associated proteins. Our drug prediction results yielded new candidates including ocriplasmin and collagenase. We also used a machine learning approach to identify the BCNHL biomarkers YES1, FERMT2, and FAM98B, novel biomarkers of high predictive fidelity. This meta-analysis validates existing knowledge while providing novel insights into the inner workings and mechanisms of B-cell lymphomas that could give rise to improved diagnostics and/or therapeutics. No external funding was used for this study.

## Introduction

Lymphomas are cancers of the blood. In 2016, there were 461,000 cases of Non-Hodgkin’s lymphoma worldwide, resulting in 240,000 deaths (1). Among non-Hodgkin’s lymphomas, only ∼10-15% are T-cell lymphomas, while the remaining 85-90% are B-cell malignancies (2). B-cell Non-Hodgkin’s Lymphomas (BCNHLs) pose a significant disease burden worldwide. BCNHL subtypes include Burkitt’s lymphoma, nodal, extra-nodal, and splenic marginal-zone B-cell lymphomas, follicular lymphoma, diffuse large B-cell lymphoma, mantle cell lymphoma, post-transplantation lymphoproliferative disorders, small lymphocytic lymphoma, lymphoblastic lymphoma, lymphoplasmacytic lymphoma, and lymphomatoid granulomatosis (2). B-cell lymphomas are dependent on their extracellular environment for activation and transformation into malignancies, including antigen activation of the B-cell receptor, canonical B-cell growth signals which are also essential to the maturation of healthy B-cells, and signals delivered by other immune cells in the follicular/germinal center lymphoma microenvironment (3).

The research community has dedicated extensive effort to identify the attributes that characterize cancers across all subtypes. Specifically, it has been suggested previously that all cancers share the following traits: selective proliferative advantage, altered stress response, vascularization, invasion and metastasis, metabolic rewiring, immune modulation, and an abetting microenvironment (4,5). One example of a molecular mechanism that is common in cancer is malignant development through TP53 mutation, with multiple mutations in the TP53 being associated with hundreds of cancer subtypes (6). Though not every gene-mechanism pairing will be found across malignant cells like TP53, it is logical to assume that identifying shared genes and mechanisms by meta-analyzing previous research in a focused set of related cancer subtypes can be beneficial. We can therefore leverage known mechanisms from well-studied subtypes to enable quicker, less expensive mechanism discovery for understudied subtypes. This approach could potentially enable researchers to develop safe and effective treatments.

The widespread adoption of RNA-sequencing (RNA-seq) has opened new frontiers in disease research. Rather than identifying and characterizing individual proteins, transcriptomic meta-analyses can provide a mechanistic snapshot of the many upregulated or downregulated genes that are affected in response to a given stimulus, such as lymphoma. Monitoring these transcriptional patterns can aid in the identification of genes that could be worth further experimental investigation due to their selective modulation in diseased samples. Though the RNA-sequencing samples in the current study were previously published, compiling them into a meta-analysis can grant us new insights into disease mechanisms by increasing the signal of significant genes and reducing the statistical “noise” caused by outliers.

The aim of this study was to identify some of the shared underlying molecular mechanisms and biomarkers of B-cell lymphomas by performing a meta-analysis of transcriptomic data from publicly available B-cell Non-Hodgkin’s Lymphomas (BCNHLs) clinical samples. We expect our analysis to validate past findings of B-cell cancer mechanisms and uncover mechanisms that have not been previously associated with BCNHL.

## Results

We acquired our BCNHL samples from the NCBI Gene Expression Omnibus (GEO) using the search term, “b-cell lymphoma” with the goal of finding B-cell non-Hodgkin’s lymphoma samples and healthy B-cell controls. We excluded non-human samples, cell lines, formalin-fixed paraffin-embedded tissues, gene expression microarray experiments, single-cell (10X) RNA-sequencing experiments, xenografts, samples known to be infected with EBV and KSHV, and samples which contained more diverse cell types (i.e., whole blood, lymph node, PBMCs, brain, etc.). We intentionally decided to not include multiple myeloma, leukemia, and Hodgkin’s lymphoma samples in favor of focusing on B-cell non-Hodgkin’s lymphomas. We then located more healthy B-cell control samples from BCNHL-unrelated studies to even out case and control numbers, the final three studies cited in Table 1. Our final dataset included a total of 322 samples (134 BCNHL samples and 188 healthy B-cell controls) from ten studies (Fig 1, Table 1, S1 File) (7–18). The samples included in our meta-analysis were all clinical samples. The risk of synthesizing study results and accounting for heterogeneity were reduced due to the lack of treatment metadata and patient outcome data as input to our analysis. Given that the aim of this study was to compare the maximum number of BCNHL samples to healthy B-cells, the only source of heterogeneity that we are concerned with is the distribution of BCNHL samples across the included subtypes. Our study was limited to GEO’s publicly available human B-cell RNA-sequencing datasets that met our criteria, and limitations may include lack of diversity in ethnicities and age groups.

**Table 1.**
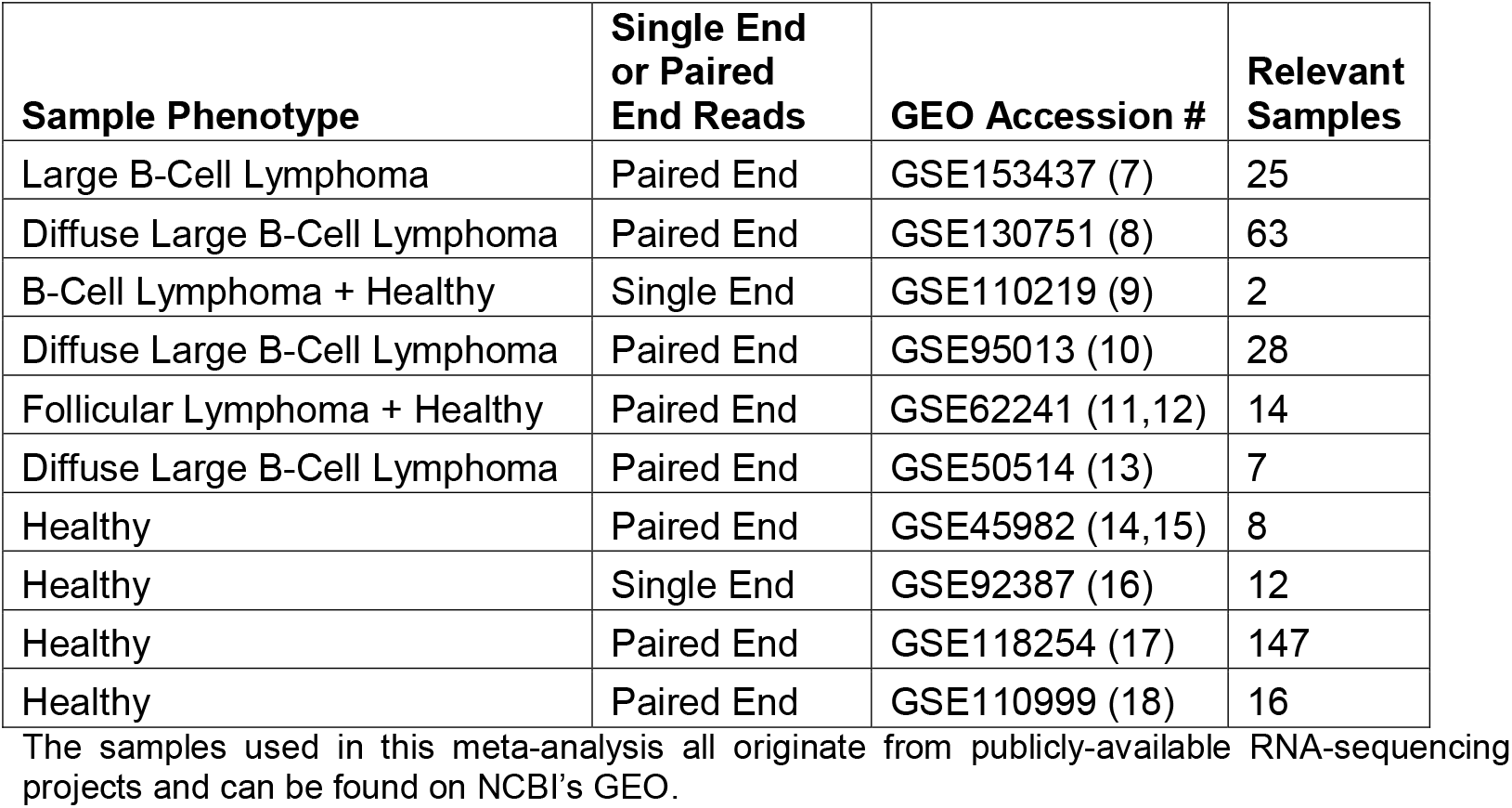
Study-based origin of samples included in the meta-analysis.

**Figure 1.**
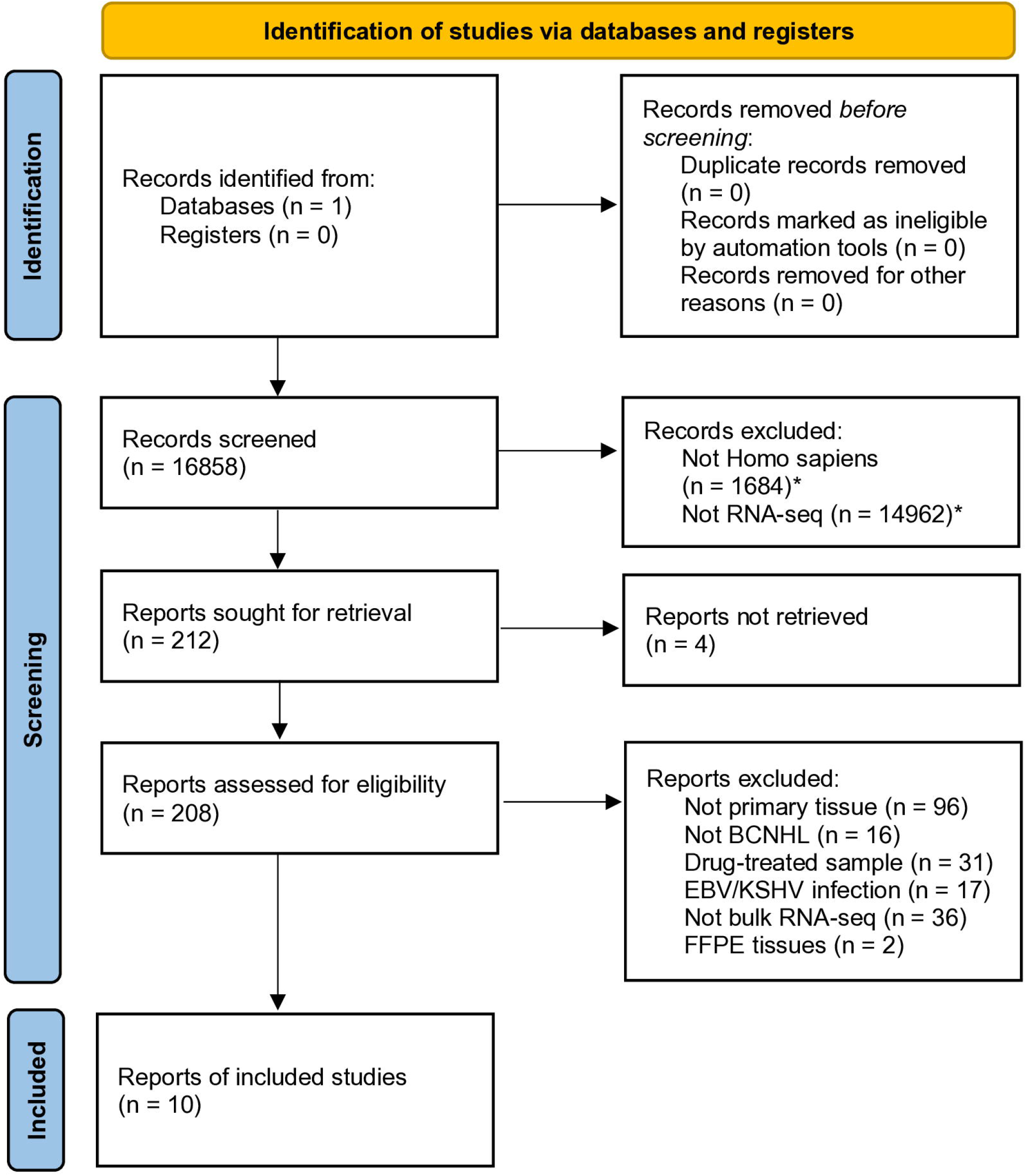
PRISMA flow diagram for transparent reporting of meta-analysis study selection. Contains a study-by-study breakdown of selection criteria. All studies included were retrieved from the Gene Expression Omnibus (GEO) database hosted by NCBI.

We began by trimming, mapping, and quantifying the reads prior to calculating the significant differential gene expression when comparing the Lymphoma to the non-diseased control samples. This comparison returned ∼13,800 significant differentially expressed genes (DEGs) (Figs 2 and 3, Table 2, S2 File). We then ranked this list by the FDR-corrected p-value for each gene. We observed that the top 20 DEGs include accepted biomarkers of various Lymphomas. Specifically, we confirmed several genes that have previously been explored or characterized in various subtypes of BCNHL including Apolipoprotein C1 (APOC1; logFC = 6.93, FDR = 8.55 × 10^−117^) and Vascular cell adhesion molecule 1 (VCAM1; logFC = 7.85, FDR = 2.29 × 10^−120^) to be upregulated in BCNHLs. We also found two pathological BCNHL genes, C-C motif chemokine ligand 18 (CCL18; logFC = 10, FDR = 3.74 × 10^−123^) and C-X-C motif chemokine ligand 9 (CXCL9; logFC = 11, FDR = 4.31 × 10^−141^) to be upregulated in BCNHL as compared to healthy B-cells.

**Table 2.**
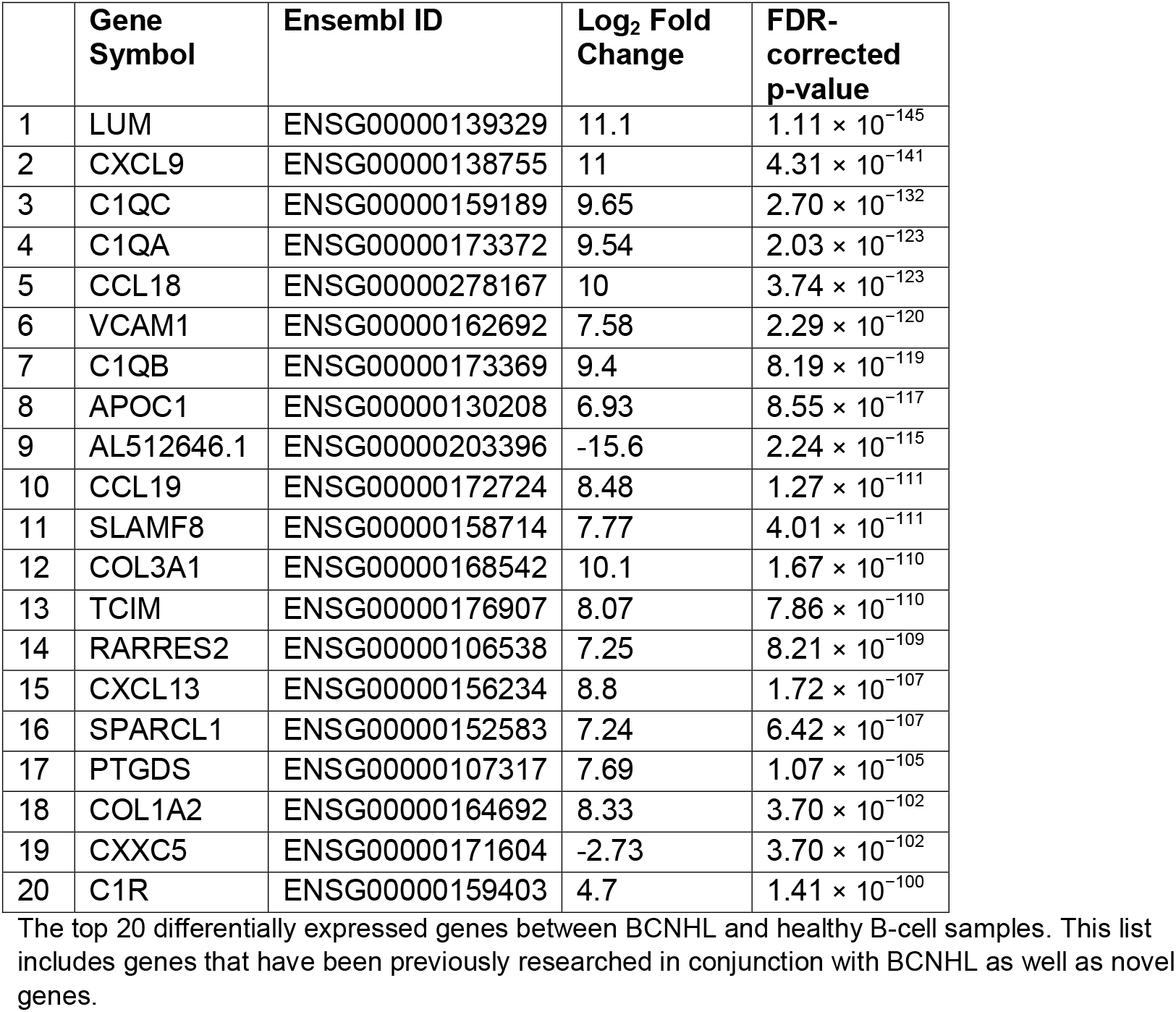
Top 20 significant differentially expressed genes.

**Figure 2.**
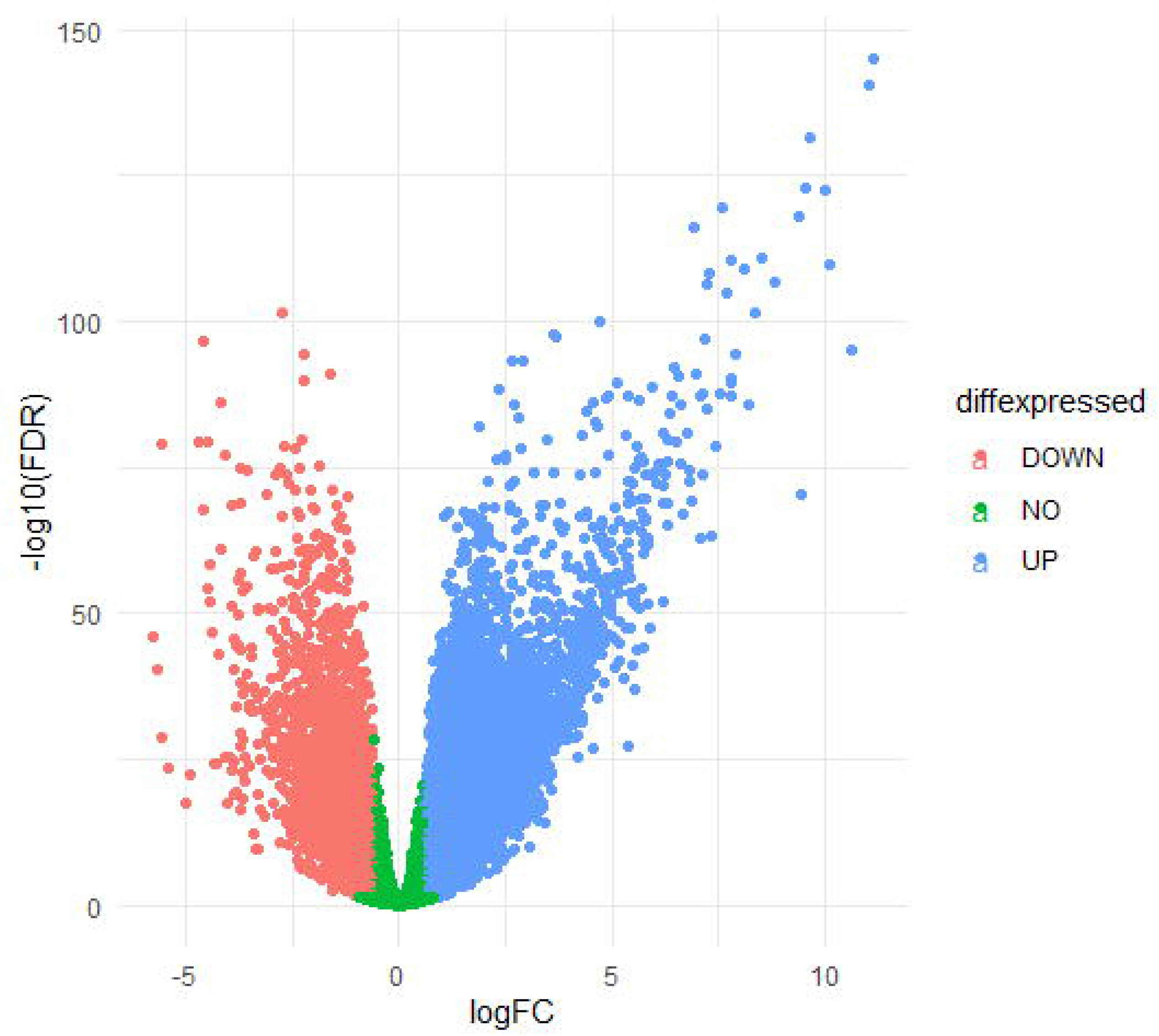
Visualization of Differentially Expressed Genes and Gene Ontologies. Differentially expressed gene volcano plot. Green dots represent genes which were not significantly differentially expressed between healthy B-cells and BCNHL, while the salmon and blue dots represent underexpressed and overexpressed genes respectively.

**Figure 3.**
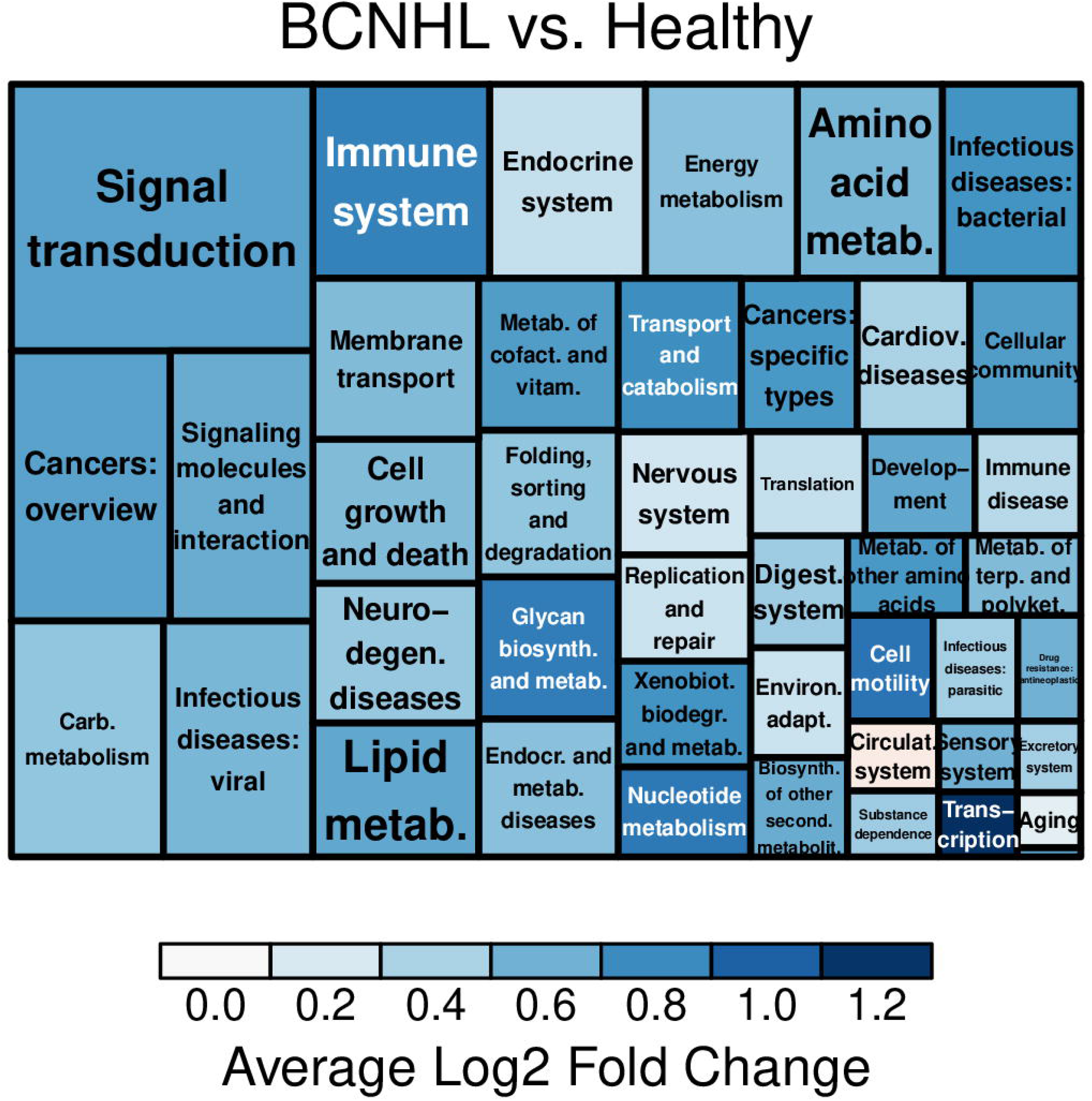
Visualization of Gene Ontology Terms. Gene ontology visualization. Each rectangle represents a gene ontology term found in the KEGG Brite gene ontology hierarchy. The size of each rectangle corresponds to the number of BCNHL differentially expressed genes present in that category. The color of each rectangle corresponds to the average log_2_ fold change of the genes included in that gene ontology. No KEGG Brite gene ontologies were found to be significantly differentially expressed by the bc3net hypergeometric enrichment.

We then examined the highest-ranking differentially expressed genes from our meta-analysis to identify gene-based mechanisms of disease. The first gene we observed using this approach was Lumican (LUM), which is a member of the small leucine-rich proteoglycans (SLRPs) (19), and was substantially upregulated in lymphoma (log_2_ fold change = 11.1, FDR p-value = 1.11 × 10^−145^). In addition, the larger family of SLRPs appears to play a role in BCNHL (Table 2). Specifically, our data show that 12/18 SLRPs are expressed in cancerous B-cells, and that 11/12 B-cell-expressed SLRPs are significantly differentially expressed in BCNHL samples. We found that overall, the SLRP fold changes substantially differed (9/12 expressed SLRPs are upregulated, 2/12 are downregulated, 1/12 had no significant change), with the genes encoding SLRPs (especially Classes I and V) being well represented in the B-cell lymphoma transcriptome.

Complement proteins are typically regarded as components of the innate immune system, which bind to antigen-antibody complexes to facilitate the formation of the membrane attack complex. We found that genes encoding the Complement C1q A (C1QA; logFC = 9.54, FDR = 2.03 × 10^−123^), Complement C1q B (C1QB; logFC = 9.4, FDR = 8.19 × 10^−119^) and Complement C1q C (C1QC; logFC = 9.65, FDR = 2.7 × 10^−132^) chains were all dramatically and significantly upregulated in BCNHL.

We detected AL512646.1 (also known as LOC100128906 and as a WDR45-like pseudogene) as differentially expressed by B-cell non-Hodgkin’s lymphoma samples, a novel observation which was somewhat unexpected. Though AL512646.1 is annotated as a pseudogene, the RNA-sequencing data shows that it is downregulated in at least a subset of BCNHLs (log_2_FC = -15.1), and it has not been previously associated with cancer.

Next, we used the DRIMSeq algorithm to determine which genes had significant differences in the presence of splice variants between case and control samples. This analysis returned 320 genes for which splice variants were significantly different (Table 4, S3 File). Apolipoprotein E (APOE) was the most statistically significant splice variant (Lr [likelihood ratio] = 4470, # of alternate splice variants = 4, adjusted p-value = 0). Specifically, we observed the expression of APOE transcripts ENST00000252486, ENST00000425718, ENST00000434152, ENST00000446996, and ENST00000485628 to significantly differ between non-Hodgkin’s lymphoma and non-diseased B-cells.

**Table 3.** Differential expression of members of the Small Leucine-Rich Proteoglycan Family (SLRPs).

| SLRP Class | Name | Log<sub>2</sub> Fold Change | FDR-corrected p-value |
| --- | --- | --- | --- |
| Class I | DCN | 2.88 | $1.67 \times 10^{-41}$ |
| | BGN | 7.88 | $3.22 \times 10^{-95}$ |
| | ASPN | 3.09 | $2.27 \times 10^{-25}$ |
| | ECM2 | 2.1 | $1.44 \times 10^{-17}$ |
|  | ECMX | NP | NP |
| Class II | FMOD | 5.71 | $3.86 \times 10^{-61}$ |
| | LUM | 11.1 | $1.11 \times 10^{-145}$ |
| | PRELP | 0.617 | $1.54 \times 10^{-4}$ |
|  | KERA | NP | NP |
|  | OMD | NP | NP |
| Class III | EPYC | NP | NP |
|  | OPTC | NP | NP |
|  | OGN | NS | NS |
| Class IV | CHAD | -3.49 | $1.33 \times 10^{-24}$ |
|  | NYX | NP | NP |
| | TSKU | 1.37 | $1.62 \times 10^{-16}$ |
| Class V | PODN | 1.66 | $5.70 \times 10^{-10}$ |
| | PODNL1 | -1.49 | $3.15 \times 10^{-11}$ |

Out of the 18 members of the SLRP family, 12 are expressed in B-cells and 11 are significantly differentially expressed between BCNHL and healthy B-cells. This is a novel finding.
\*NS = not significant; NP = not present.

**Table 4.**
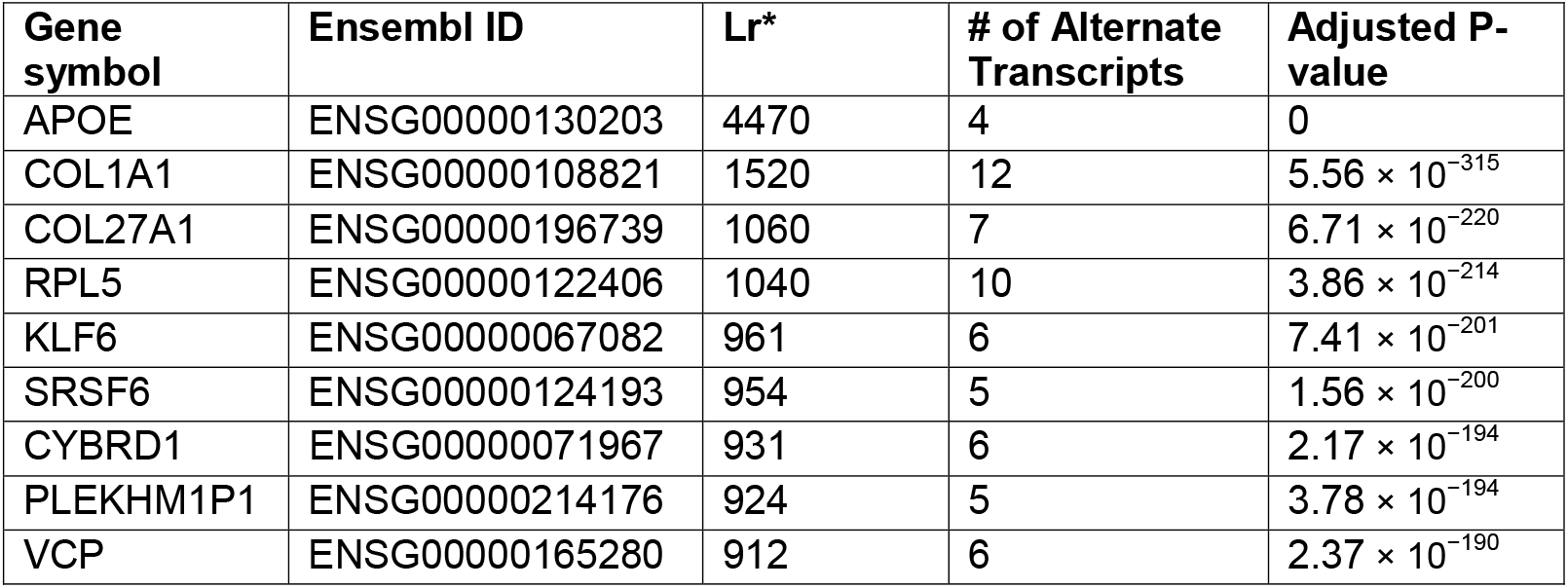
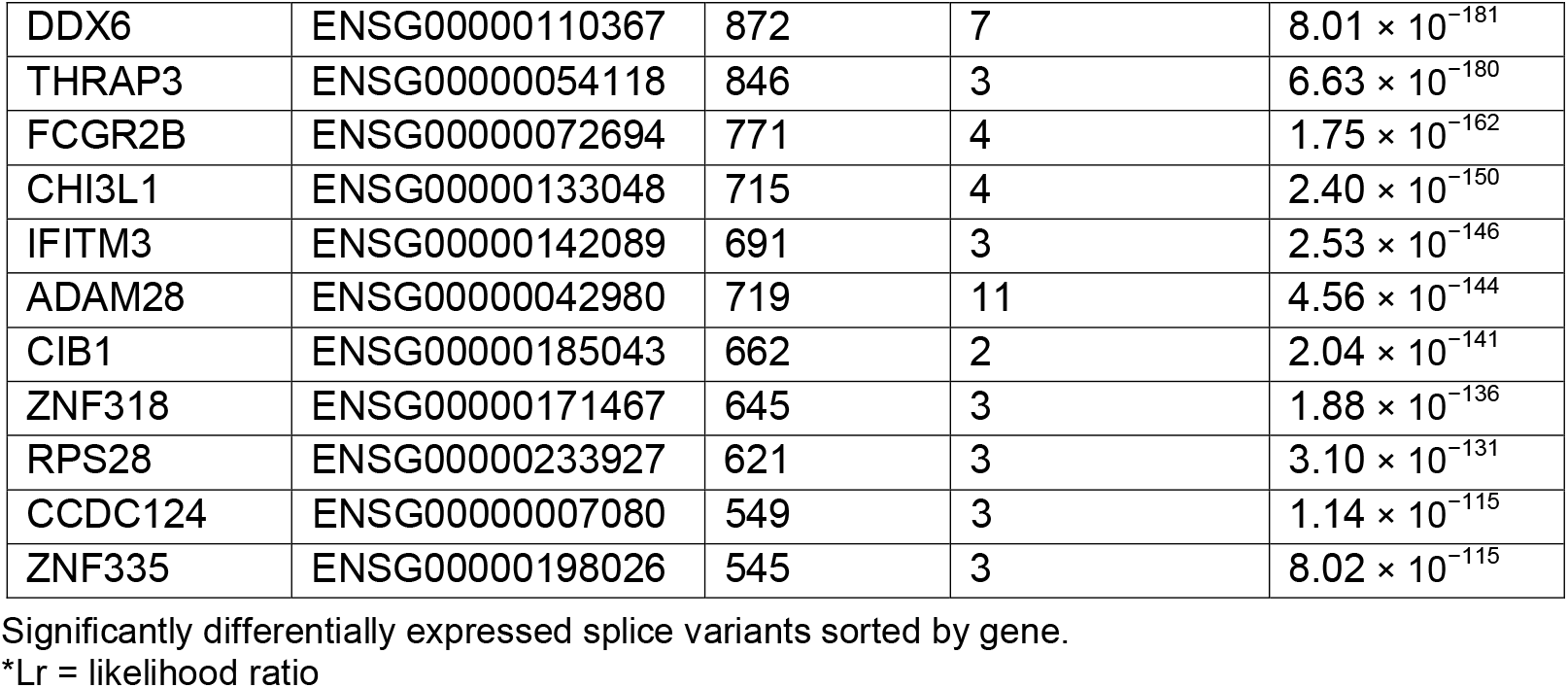
Top 20 most significant splice variants (by gene).

We also observed that Collagen type I alpha 1 chain (COL1A1) had significant splice variants (Lr = 1520, # of alternate splice variants = 12, adjusted p-value = 5.55999999807983 × 10^−315^). Interestingly, our study also found that the COL1A1 gene was significantly upregulated in BCNHL (logFC = 3.73, FDR = 9.78 × 10^−48^). We also observed novel significant splice variants in Collagen type XXVII alpha 1 chain (COL27A1), which was found to be significant in BCNHL (Lr = 1060, # of alternate splice variants = 7, adjusted p-value = 6.71 × 10^−220^).

We then wanted to determine which functional terms in the Gene Ontology were over-represented by the list of DEGs in BCNHL. The Camera algorithm checked 14,901 terms (including gene ontologies and human phenotypes) for enrichment against the significant differentially expressed genes that we generated with edgeR. Although there were 482 results (p-value < 0.05), none remained significant after multiple hypothesis correction (S4 File). The lack of significant results is somewhat expected given the overall molecular heterogeneity of BCNHL subtypes. To visualize the gene ontology changes, we used a hypergeometric enrichment algorithm that applied a p-value cutoff of 0.05. We then averaged the edgeR fold-change values for the genes of each gene ontology in the KEGG Brite hierarchy and plotted the enrichment results using the R Treemap package to better understand the contribution of various terms to the overall list of DEGs (Fig 3).

To better understand the results of our analysis at a more mechanistic level, we used the signaling pathway impact analysis (SPIA) algorithm to identify intracellular signaling pathways that play important roles in Lymphoma. This pathway analysis generates a null distribution through bootstrapping to identify pathways that are significantly modulated when comparing sets of samples. Our analysis revealed 33 significantly modulated pathways between lymphoma B-cells and non-diseased B-cells (Table 5, S5 File). Specifically, we observed eight pathways that were involved with the extracellular matrix and connective tissue in general, including Integrin signaling pathway, Extracellular matrix organization, ECM-receptor interaction, Focal adhesion, Integrins in angiogenesis, integrin signaling pathway, Collagen formation, and Collagen degradation. The upregulation of these pathways indicate that BCNHL likely benefits from modulations to the extracellular matrix.

**Table 5.** Significant differentially modulated signaling pathways.

|  | Name | pSize | NDE | tA | pGFWER | Source Database |
| --- | --- | --- | --- | --- | --- | --- |
| 1 | Integrin signalling pathway | 99 | 86 | 114.394 | $2.39 \times 10^{-5}$ | Panther |
| 2 | Extracellular matrix organization | 204 | 180 | 80.7398395 | $3.14 \times 10^{-5}$ | Reactome |
| 3 | ECM-receptor interaction | 70 | 64 | 77.659 | $4.47 \times 10^{-5}$ | KEGG |
| 4 | Staphylococcus aureus infection | 32 | 29 | 110.568396 | 0.000503428 | KEGG |
| 5 | Complement and coagulation cascades | 36 | 32 | 37.5461944 | 0.000674242 | KEGG |
| 6 | Urokinase-type plasminogen activator (uPA) and uPAR-mediated signaling | 28 | 25 | 130.821315 | 0.000740169 | NCI |
| 7 | Cytokine-cytokine receptor interaction | 168 | 140 | 98.294 | 0.000780995 | KEGG |
| 8 | Focal adhesion | 182 | 150 | 236.615459 | 0.001133449 | KEGG |
| 9 | PI3K-Akt signaling pathway | 271 | 221 | 260.046197 | 0.001344307 | KEGG |
| 10 | Complement cascade | 29 | 27 | 102.278583 | 0.001440498 | Reactome |
| 11 | Systemic lupus erythematosus | 17 | 15 | 67.4577222 | 0.001616635 | KEGG |
| 12 | b cell survival pathway | 22 | 19 | 26.576 | 0.00167109 | BioCarta |
| 13 | Small cell lung cancer | 78 | 64 | 121.170067 | 0.001930414 | KEGG |
| 14 | Integrins in angiogenesis | 52 | 41 | 146.143424 | 0.00285984 | NCI |
| 15 | Olfactory transduction | 93 | 74 | -148.8965 | 0.002966626 | KEGG |
| 16 | integrin signaling pathway | 37 | 29 | 77.0156667 | 0.003336442 | BioCarta |
| 17 | erk and pi-3 kinase are necessary for collagen binding in corneal epithelia | 34 | 26 | 166.268917 | 0.003755165 | BioCarta |
| 18 | RNA Polymerase I Promoter Clearance | 85 | 72 | -40.156 | 0.004307328 | Reactome |
| 19 | Initial triggering of complement | 15 | 14 | 44.508 | 0.004480759 | Reactome |
| 20 | RNA Polymerase I Promoter Opening | 39 | 34 | -40.907 | 0.004675938 | Reactome |
| 21 | RHO GTPases activate PKNs | 67 | 57 | 39.779 | 0.004802382 | Reactome |
| 22 | DNA Damage/Telomere Stress Induced Senescence | 61 | 52 | 32.7708077 | 0.004980633 | Reactome |
| 23 | Creation of C4 and C2 activators | 7 | 7 | 27.365 | 0.005633091 | Reactome |
| 24 | Collagen formation | 66 | 63 | 26.4272897 | 0.006045837 | Reactome |
| 25 | Activated PKN1 stimulates transcription of AR (androgen receptor) regulated genes KLK2 and KLK3 | 41 | 35 | 39.094 | 0.006675281 | Reactome |
| 26 | MET activates PTK2 signaling | 18 | 16 | 63.573 | 0.007079287 | Reactome |
| 27 | Collagen degradation | 17 | 15 | 131.8905 | 0.008037505 | Reactome |
| 28 | MET promotes cell motility | 28 | 24 | 97.5445 | 0.00808298 | Reactome |
| 29 | Regulation of IGF Activity by IGFBP | 11 | 10 | 25.958725 | 0.008404989 | Reactome |
| 30 | Classical antibody-mediated complement activation | 5 | 5 | 27.354 | 0.008619298 | Reactome |
| 31 | Serotonin Neurotransmitter Release Cycle | 11 | 9 | -13.301889 | 0.015608196 | Reactome |
| 32 | Class A/1 (Rhodopsin-like receptors) | 81 | 77 | 5.908 | 0.026176599 | Reactome |
| 33 | Peptide ligand-binding receptors | 79 | 75 | 5.84 | 0.040928099 | Reactome |

The significantly differentially modulated pathway results. Included are nine extracellular matrix-associated pathways.
\*Abbreviations: psize = number of genes in pathway. NDE = number of genes from pathway which were differentially expressed. tA = measure of change between healthy and lymphoma expression; directionality indicates up- or down-regulation. pGFWER = p-value with adjustments appropriate to a multiplexed interaction network. (20)

We next used the Pathways2Targets algorithm to identify potentially novel drug targets for BCNHL from the signaling pathway results (S6 File). We sorted the results so that drug targets present in multiple signaling pathways would be ranked higher (Table 6, S7 File). We predicted the most relevant existing FDA-approved drugs for other indications that could affect the lymphoma phenotype are Doxycycline, Ocriplasmin, and Collagenase. We also identified ATN-161 as a candidate drug, but it has only been tested in phase-two trials.

**Table 6.**
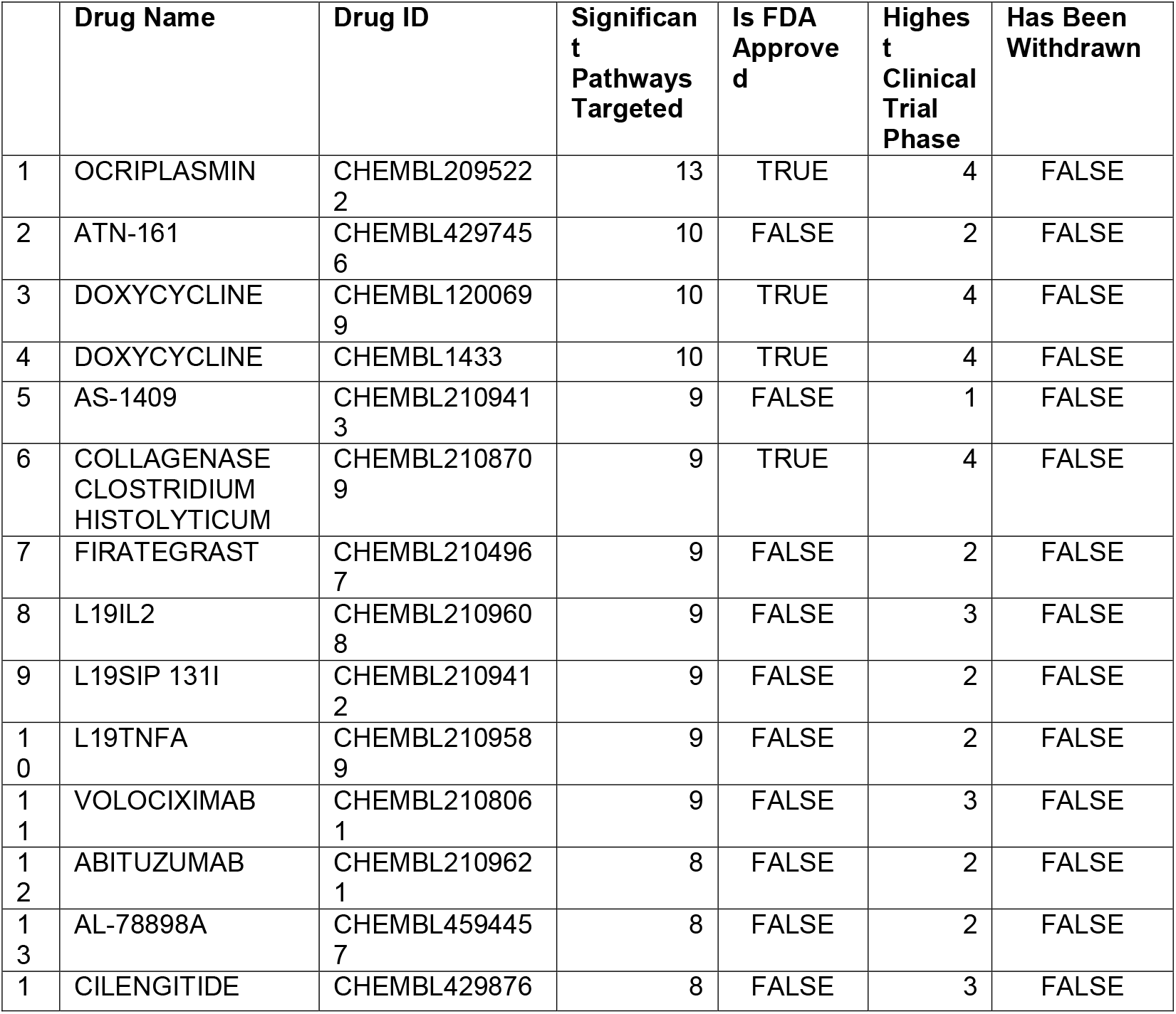
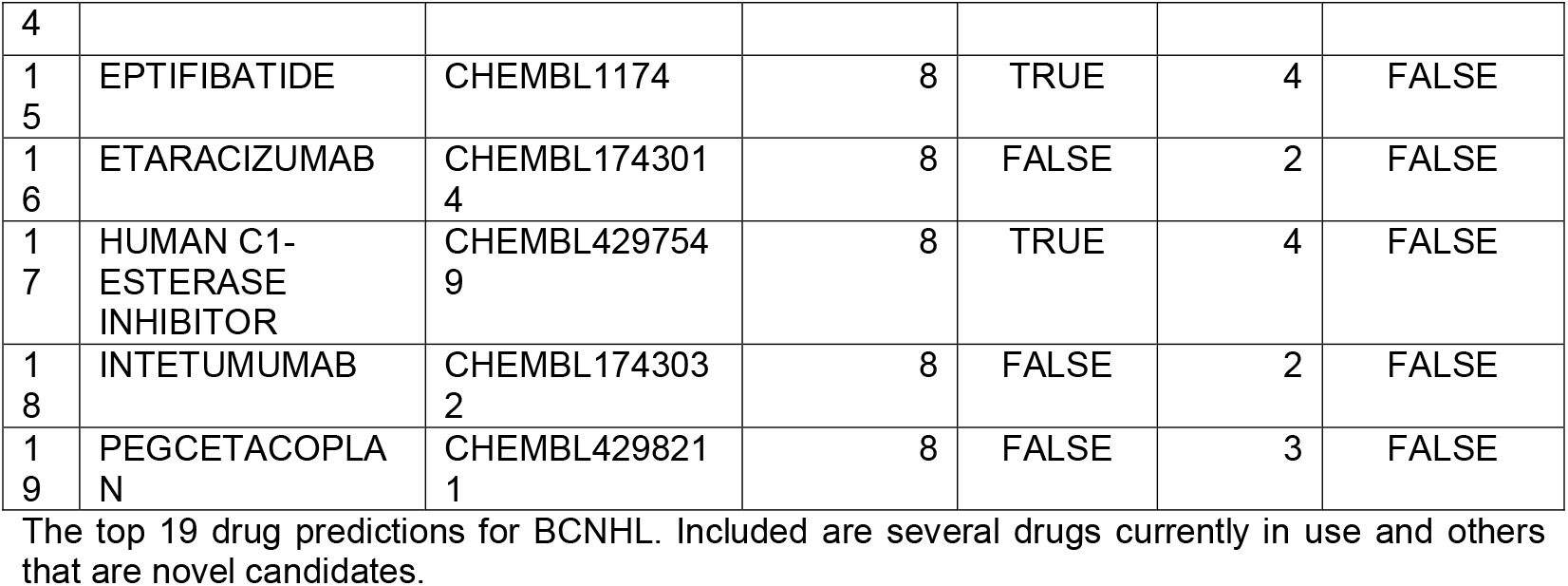
Predicted BCNHL drugs based on signaling pathways.

Rather than solely rely on the significant differential expression data to determine biomarkers, we applied a more robust random forest machine learning method to predict biomarkers of BCNHLs. Specifically, the DEG statistics focus on identifying genes that have a large difference in expression between two states, while the random forest approach identifies genes that consistently change across disease vs. healthy samples. Consequently, the random forest approach identifies transcripts that are best capable of differentiating between disease and healthy states. The top three genes identified by our random forest analysis included YES1, FAM98B, and FERMT2 (Table 7, Figs 4A and 4B, S8 File). We then calculated the area under the curve for the receiver-operator characteristic curve, which showed that when the expression values from these three genes are combined they are 99.889% accurate at predicting whether the patient samples had BCNHL (Fig 4C).

**Table 7.** BCNHL biomarkers predicted from gene expression using machine learning.

| Gene Symbol | Mean Gini Decrease | edgeR Log<sub>2</sub> Fold Change | edgeR False Discovery Rate | Disease Status | Mean (Read Counts) | Standard Deviation (Read Counts) | Median (Read Counts) |
| --- | --- | --- | --- | --- | --- | --- | --- |
| YES1 | 0.77 | 2.38 | 1.98x10<sup>-38</sup> | Lymphoma | 1151.756 | 1246.946 | 629 |
|  |  |  |  | Healthy | 38.87234 | 66.01043 | 11 |
| FAM98B | 0.68 | 1.58 | 1.48x10<sup>-60</sup> | Lymphoma | 1797.452 | 1174.797 | 1456 |
|  |  |  |  | Healthy | 248.4202 | 954.1375 | 32 |
| FERMT2 | 0.67 | 2.83 | 1.46x10<sup>-38</sup> | Lymphoma | 1246.993 | 1200.669 | 841 |
|  |  |  |  | Healthy | 32.46809 | 73.10299 | 4 |

The top three genes identified by our random forest biomarker prediction are high-fidelity biomarkers of BCNHL due to their consistent and extreme upregulation across our 134 clinical BCNHL samples as compared to our 188 healthy B-cell samples. Presented above are statistics that capture the spread of transcriptional levels between BCNHL and healthy groups.

**Figure 4.**
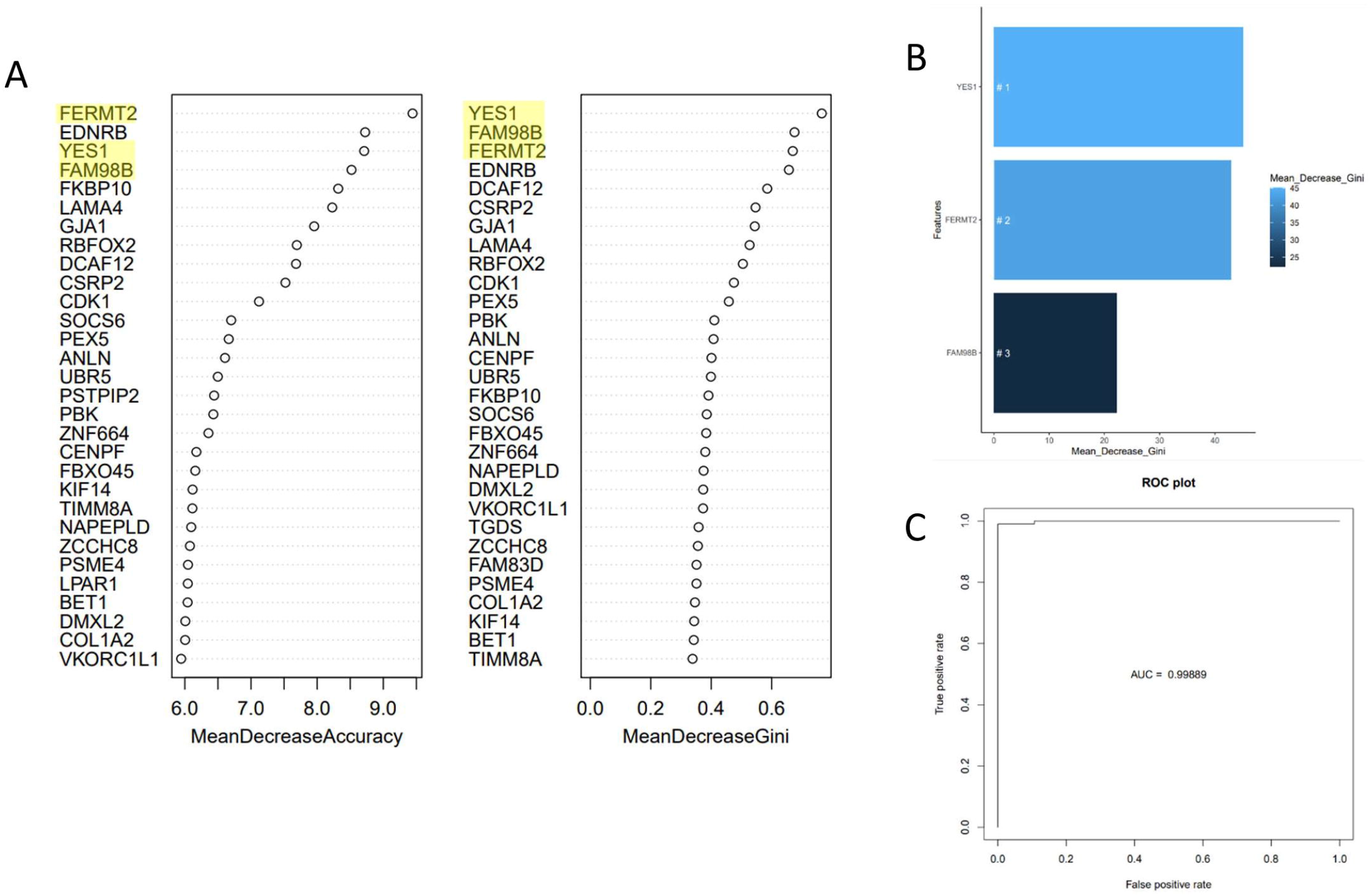
Biomarker Prediction Yields Three-Gene Signature with 99% Predictive Ability. A) Biomarkers ranked by mean decrease in Gini impurity and permutation values using random forest show YES1, FAM98B, and FERMT2 as the highest ranked predictive biomarkers (ranked by mean decrease of Gini impurity score). B) Random forest biomarker prediction for the top three genes in isolation. C) Receiver-operator characteristic curve using only YES1, FAM98B, and FERMT2 shows these three genes are 99.889% accurate at predicting BCNHL status (healthy or diseased).

## Discussion

The goal of this study was to collect publicly available RNA-seq data from GEO and process those data to find differentially expressed genes, pathways, splice variants, and biomarkers. We confirmed several biologically- and clinically relevant biomarkers and pathologic mechanisms that were identified previously, as well as novel entities. We found several key genes that are significantly differentially expressed in BCNHL including LUM and other SLRPs, complement protein components, and the supposed pseudogene AL512646.1. We confirmed that previously characterized biomarkers such as APOC1, VCAM1, CCL18, and CXCL9 are overexpressed in BCNHL, and that 320 genes including APOE, COL1A1, and COL27A1 had differentially expressed splice variants. We additionally found a BCNHL reliance on the upregulation of pathways associated with the extracellular matrix.

To our knowledge, this is the largest meta-analysis of human samples in the BCNHL field to-date. While some may be concerned that the signals from the individual subtypes of non-Hodgkin’s B-cell lymphomas could drown one another out, we believe that including representative samples from multiple BCNHL subtypes augments the signal(s) that are shared among the represented subtypes and could aid in the identification of shared mechanistic insights with reduced bias. One potential limitation of our study was to only evaluate samples from the GEO database for inclusion in our analysis. Our intentional focus on BCNHL excluded multiple myelomas, B-cell leukemias, or Hodgkin’s B-cell lymphomas. Promising future directions may include mining additional public databases for similar data and potentially expanding the scope of any future meta-analysis to include all B-cell malignancies. In addition, it is possible that our focus on incorporating multiple publicly available datasets may have introduced potential biases in patient age, gender, or ethnicity.

Though there is evidence in the literature that directly associate BCNHL to some of our results (e.g. genes, splice variants, and pathways), some of our findings are novel to BCNHL. In cases where no previously published research indicates the relationship between BCNHL and our results, we will appeal to the Hallmarks of Cancer to investigate the relationship between our differentially expressed result and a distinct cancer system (4). We therefore ranked both the accuracy and confidence in our results by their relevance to BCNHL, followed by B-cell cancers in general, all blood cancers, and finally all cancers. We believe that identifying a possible mechanism for a gene that is associated with other cancers, and unresearched in BCNHL is still relevant. We expect that a subset of these findings will justify additional wet lab experimentation.

### Differentially expressed genes suggest shared underlying mechanisms for lymphomas

LUM seems to play a role in the progression or non-progression of several different cancer types. Mahadevan *et al*. previously reported upregulated LUM in both T- and B-cell lymphomas, but offered no insights on potential mechanisms (21). A literature search of parallel systems revealed that in breast cancer, high stromal-cell expression of LUM adjacent to the tumor stalls tumor growth, and lowered stromal expression of LUM correlates with higher breast cancer mortality rates and increased severity (22). In melanoma, LUM in the extracellular matrix halts metastasis through direct interaction with alpha-2-beta-1 integrin (23). Both breast cancer and pancreatic cancer cells have been documented to upregulate LUM, along with many other cancer types (19). Overall, LUM expression by cancer cells seems to correlate with more aggressive cancers and poorer patient outcomes. The massive LUM upregulation illustrated in our samples may be due to the fact that the BCNHL samples available on GEO were mostly from advanced or refractory cases of BCNHL. The prior finding that high LUM expression around tumors is protective against metastasis in several cancer subtypes indicates the potential for LUM as a cancer-stalling therapy.

Interestingly, a subset of the members in the SLRP protein family have been previously identified in B-cell Non-Hodgkin’s lymphomas including DCN (24), BGN (24), ASPN (25), FMOD (26), LUM (21), PRELP (26), and TSKU (27). However, other members within the SLRP family have not been previously considered as lymphoma biomarkers or potential pathology-inducing molecules. Our novel finding is that the SLRPs ECM2, CHAD, PODN, and PODNL1 are differentially expressed in BCNHL. Proteoglycans have been shown to be associated with pro-cancer mechanisms in prostate, breast, colon, lung, ovary, mesothelium, pancreatic, lymphoma, and esophageal cancers (19). Our results show two upregulated pathways in BCNHL that were previously shown to be mechanistically intertwined with proteoglycans in cancer, which are the Focal Adhesion pathway (28) and the PI3K-Akt signaling pathway (29). Taken together, this may suggest a connection between previously established proteoglycan cancer mechanisms and B-cell non-Hodgkin’s lymphomas. Additional work is still required to elucidate the role(s) that these entities play in BCNHL.

In the context of other cancers, increased expression of complement genes C1QA and C1QB at week 16 of mantle cell lymphoma treatment by Venetoclax and Ibrutinib was significantly associated with a worse prognosis (30), illustrating that C1QA and C1QB may be associated with resistance to cancer drugs. Jiang et al. showed via immunohistochemistry that C1QB localizes to the nuclei of gastric cancer cells (31). C1QB’s nuclear localization suggests that C1QB may have additional function(s). Upregulation of C1QA, C1QB, and C1QC in peripheral T-cell lymphoma (32) and upregulation of C1QC in Epstein-Barr Virus-positive diffuse large B-cell lymphoma (33) have been reported previously. In other in-vitro and in-vivo cancer models, the whole C1q protein has been shown to mediate metastasis, motility, growth and proliferation, and adhesion (34). Our results add to the growing body of work suggesting a potential alternate function of complement proteins in cancer that warrants further investigation.

In addition to our novel findings on differentially expressed genes, we were also able to detect statistically significant genes that were previously characterized in at least one subtype of BCNHL. The first of these proteins is Apolipoprotein C1 (APOC1), which we observed to be upregulated in BCNHL. APOC1 is one of three genes whose expression levels are predictive of diffuse large B-cell lymphoma severity (35), and it is also upregulated in late stage lung cancers as compared to early stage lung cancers (36). This suggests that APOC1 may be contributing to cancer pathology across diverse cancers in multiple cell types.

Our observation that C-C motif chemokine ligand 18 (CCL18), which has a well-recognized role in lymphoma, was upregulated in our BCNHL analysis is relevant since this gene assists large B-cell lymphoma in cell proliferation, the NF-Kappa-B pathway, and the PI3K-AKT pathway (37). Its upregulation in macrophages and dendritic cells from cutaneous T-cell lymphoma lesions was associated with a negative prognosis (38).

Our finding C-X-C motif chemokine ligand 9 (CXCL9) to be significantly upregulated in our analysis of B-cells is interesting since this gene has been shown to promote the progression of diffuse large B-cell lymphoma by halting degradation of beta-catenin (CTNNB1) and upregulating its initial expression (39). Our findings support this proposed mechanism with CTNNB1 being upregulated in lymphoma (log_2_FC = 1.1, FDR = 1.54 × 10^−33^), while other elements of the CTNNB1 “destruction complex” were mostly downregulated. Specifically, several of the known components of the destruction complex that were detected in our analysis include APC (log_2_FC = -0.755, FDR = 3.51 × 10^−11^), GSK3B (log_2_FC = -0.692, FDR = 2.62 × 10^−3^), CSNK1A1 (not significant), AXIN1 (log_2_FC = 0.533, FDR = 3.96 × 10^−10^), BTRC (not significant), and FBW11 (log_2_FC = -0.692, FDR = 5.60 × 10^−20^).

We identified several other genes of that may be relevant to pathogenesis. Small but significant upregulation of AXIN1 is of interest for additional investigation due to its ties to CXCL9, and is not known to have multiple heterogenous functions (40). AXIN1 regulates the Wnt and JNK signaling pathways (41), and it regulates the Wnt pathway by degrading CTNNB1 (39). If CTNNB1 isn’t degraded by AXIN1, CTNNB1 translocates to the nucleus and interacts with LEF1, which we found to be significantly upregulated, and TCF7 (not significant in this study), causing transcription of Wnt pathway target genes to occur (42,43). Wnt helps to regulate cell cycle and contributes to the increased growth rate of many cancer types (44). AXIN1 activates the JNK signaling pathway by binding to MAP3K1, which we found to be significantly downregulated, or to MAP3K4, which was significantly upregulated (45). Since CTNNB1 has been shown to contribute to apoptosis resistance in multiple myeloma cells (46), it is possible that the inability to stop the destruction of CTNNB1 in lymphoma may share a similar mechanism.

Finally, VCAM1 upregulation is associated with a poor prognosis for patients with non-Hodgkin’s lymphomas, and VCAM1 is under investigation as a potential serum biomarker for assessing disease progression (47). Adhesion molecules such as VCAM1 promote cancer metastasis, or in the case of blood cancers, extravasation, by allowing cancer cells to exit the bloodstream and integrate with healthy tissues throughout the body (48).

### Splice variants suggest relevance to lymphomas

To better understand the contribution of differentially expressed splice variants to disease, we examined the highest-ranked DRIMseq results. This algorithm calculates statistical significance based on the number of reads mapped to exons that are present in each splice variant. Our observation that Apolipoprotein E (APOE) was the highest-ranking splice variant result validates previous findings that associate this gene with pancreatic cancer pathology (49). In addition, pediatric patients with malignant lymphoma and acute lymphoblastic leukemia who express isoforms E3 and E4 of APOE are at higher risk of developing extreme hypertriglyceridemia (50). Though little research has been done concerning the mechanisms of APOE in BCNHL, we believe that APOE may be contributing to disease by participating in the Regulation of Insulin-like Growth Factor (IGF) activity by Insulin-like Growth Factor Binding Protein (IGFBP) pathway, which is we found to be a significantly modulated pathway that includes APOE. The significance of APOC1 as a DEG in BCNHL, paired with the evidence of significant APOE splice variants suggest that apolipoproteins may be useful targets for future BCNHL treatments.

Our observation of Collagen type I alpha 1 chain (COL1A1) as a highly ranked splice variant result is novel to the best of our knowledge. However, the literature indicates that the COLA1A-014 transcript regulates the CXCL12-CXCR4 axis in gastric cancer, leading to tumor progression (46). In addition to displaying significant differences in splice variant expression, we also found COL1A1 to be significantly upregulated in BCNHL. COL1A1 has previously been reported to be upregulated in peripheral T-cell lymphoma (21). In Hodgkin’s lymphoma, COL1A1 overexpression is associated with epigenetic silencing of the RNA demethylase ALKBH3 and reduced survival (51). COL1A1 is a member of several of our significant upregulated pathways involving the extracellular matrix (ECM-receptor interaction, Focal adhesion, Extracellular matrix organization, and Collagen formation). This involvement in extracellular matrix-related pathways strengthens the case that the mechanism of COL1A1 may involve tumor cell interaction with its outer environment.

Collagen type XXVII alpha 1 chain (COL27A1) having significant changes among its expressed splice variants in BCNHL is interesting since it was recently reported as being overexpressed in adenoid cystic carcinoma (52). Like COL1A1, COL27A1 is a member of the upregulated Extracellular matrix organization and Collagen formation pathways, suggesting that COL27A1 could play a role in BCNHL extravasation.

### Extracellular matrix-related pathways may contribute to disease

Our signaling pathway enrichment analysis broadened the scope of our analysis and interpretation. Many of our findings supported an interesting reliance of BCNHL on pathways associated with the extracellular matrix. Recent research has suggested the importance of extracellular matrix components in reactivating quiescent cancer cells through the β1-integrin signaling pathway (53). It would follow that interaction with extracellular matrix components also plays a role in regulating cancer cells. To our knowledge, no studies have reported the integrin signaling pathway to be activated in BCNHL, though it has been reported as activated in the closely-related cancer NK/T-cell lymphoma (21). The activation of these pathways suggests that malignant BCNHL cells may have an advantage by interacting with the extracellular matrix. Such interactions with the extracellular matrix are typically considered to be an important part of metastasis (48). We found this result to be interesting since lymphomas are liquid tumors, unbound by extracellular matrix. This upregulation of pathways allowing interaction with the extracellular matrix may suggest that BCNHL could be invading non-lymphatic and/or non-circulatory tissues.

The trend of extracellular matrix interaction is also seen in the DEG results, adding support to the idea that interaction with the extracellular matrix is important for BCNHL growth and survival. Additionally, COL1A1 and COL27A1, which are members of extracellular matrix-related pathways, are two of the genes with the most significantly differential expression of splice variants.

### Drug prediction algorithm returned both tested and novel candidates

Of our top drug results, doxycycline is currently in use for ocular B-cell lymphomas (54,55). It is additionally under investigation for diffuse large B-cell lymphoma; recent work found doxycycline suppresses diffuse large B-cell lymphoma growth *in vitro* and *in vivo* via CSN5 inhibition (56). ATN-161 is a novel drug candidate for BCNHL. Though it is only in phase two of clinical trials, it has been a successful drug against refractory solid tumors, making it a promising drug candidate for other susceptible malignancies (57). ATN-161 suppresses cancer via integrin beta1 alpha5 antagonism, disabling invasion and metastasis (58). Ocriplasmin reverses vitreomacular adhesion via interaction with fibronectin and laminin (59). Though ocriplasmin has never been used in cancer before, it may be a promising drug candidate due to its ability to modulate adhesion. Collagenase clostridium histolyticum is under investigation for treating collagen-rich uterine fibroids and was successful at reducing the stiffness of the tumors (60).

### Machine learning predicts novel biomarkers of BCNHL

YES1, FERMT2, and FAM98B are novel biomarkers not previously associated with BCNHL. However, each has well-documented cancer associations. YES1 is a tyrosine kinase which regulates cell cycle and apoptosis *in vitro* and cell growth *in vivo* of tumors with YES1 amplification (61). YES1 has been previously identified as a biomarker for non-small cell lung cancer and esophageal adenocarcinoma (62,63) and may be a potential membrane biomarker. YES1 can anchor to the inner membrane with help from peptide SMIM30 (64), but whether it can flip to the outer leaflet has not been investigated. The role of YES1 in BCNHL pathogenesis also needs additional investigation. FERMT2 has previously been pinpointed as a biomarker for other cancers previously including non-small cell lung cancer and prostate cancer (65,66), but not for BCNHL. FERMT2 stabilizes CTNNB1, which is a well-documented activator of oncogene transcription, and is implicated in Wnt pathway regulation (67). Additionally, FERMT2 enhances integrin signaling and mediates migration, invasion, and focal adhesion (68,69). Though FAM98B has been shown to play an important role in the development of multiple cancers, it has not previously been identified as a biomarker for any cancer. FAM98B is an arginine methyltransferase, is utilized in tumorigenesis, and works in tandem with DDX1, a pan-cancer marker, in RNA metabolism/processing (70,71). Like YES1 and FERMT2, FAM98B has not been previously identified as a biomarker for BCNHL. These three genes have substantial diagnostic potential as a liquid biopsy that could be generalizable across B-cell non-Hodgkin’s lymphoma subtypes. Further validation is needed to determine whether these are suitable biomarkers for diagnostic or screening application.

In summary, our meta-analysis identified many significant DEGs and pathways that play a role in B-cell non-Hodgkin’s lymphomas. Our findings confirm results of previous BCNHL research, indicating that the statistical analyses applied within our computational workflow pipeline are effective at accurately identifying statistically significant genes, splice variants, and pathways with clinical and pathological relevance. Additionally, several of our results are novel, which need additional validation in future experiments. It is likely that at least some of these novel findings were detected due to the ability of our meta-analysis to reduce the statistical “noise” produced by outliers from individual studies and increase the biologically-relevant signal. Specifically, our findings suggest that LUM and 10 other small leucine-rich proteoglycans are significantly differentially expressed in BCNHL, that AL512646.1 is not a pseudo-gene, that APOE, COL1A1, and COL27a1 have significant differentially expressed splice variants in BCNHL, and that BCNHL is strongly reliant on the overexpression of extracellular matrix-associated pathways. The predominant drug prediction results nearly universally targeted extracellular matrix-associated mechanisms, and has yielded several promising new potential drug candidates including ocriplasmin and ATN-161. Our random forest biomarker discovery pinpointed three novel biomarker genes not previously associated with BCNHL, YES1, FERMT2, and FAM98B, which show high fidelity in predicting lymphoma presence based on transcriptional levels. These findings shed additional light on the underlying intracellular mechanisms of BCNHL and could be used in the development of improved diagnostics and therapeutics to further improve human health.

## Methods

### Collecting samples

RNA-sequencing samples were acquired from the National Center for Biotechnology Information (NCBI) Gene Expression Omnibus (GEO) database using the search term, “b-cell lymphoma” to find B-cell non-Hodgkin’s lymphoma samples and healthy B-cell controls. The automatic GEO filters “Homo sapiens” and “high-throughput RNA-sequencing” were applied. Cell lines, formalin-fixed paraffin-embedded tissues, gene expression microarray experiments, single-cell (10X) RNA-sequencing experiments, xenografts, samples known to be infected with EBV and KSHV, and samples which contained more diverse cell types (i.e., whole blood, lymph node, PBMCs, brain, etc.) were manually excluded. All samples that had one or more of these disqualifying attributes were excluded from the dataset prior to analysis, which may have included only a subset of samples from any individual study in the meta-analysis. Multiple myeloma, leukemia, and Hodgkin’s lymphoma samples were intentionally excluded in favor of focusing on B-cell non-Hodgkin’s lymphomas. Records were accepted or rejected based on the standardized exclusion criteria detailed above by our team. To avoid inclusion bias, any sample that could not be excluded by the standardized exclusion criteria was included in the study. While a subset of the healthy control samples was obtained from the same RNA sequencing projects as the BCNHL samples, others were obtained from three unrelated B-cell datasets with healthy controls to create roughly equivalent-sized BCNHL and healthy groups. Final dataset assembly from GEO concluded on October 22, 2020, resulting in a dataset of 322 samples (134 BCNHL samples and 188 healthy B-cell controls) from ten studies (7–18). The raw data for these experiments were previously collected by the data providers and conform to the appropriate ethical oversight to protect patient autonomy and patient identity. All 10 primary RNA-sequencing datasets from which samples were gathered for our lymphoma meta-analysis have been published in the peer-reviewed literature, increasing overall confidence that each dataset has acceptable quality (Table 1, Fig 1).

### Preprocessing of RNA-sequencing data

Following the manual curation of the RNA-seq samples, the fastq files were pre-processed as previously described (72). In brief, fastq files containing RNA-sequencing data were downloaded from the Sequence Read Archive (SRA) using the sratools software package. The fastq files, the associated metadata file, and a configuration file for each dataset were then generated and used as input to the Automated Reproducible MOdular Workflow for Preprocessing and Differential Analysis of RNA-seq Data (ARMOR) workflow (73). A configuration file was used by ARMOR to appropriately set up a python-based Snakemake workflow (74). In the ARMOR workflow, adapters and poor-quality regions of reads were trimmed with TrimGalore! (75), quality control metrics were calculated with FastQC (76), reads were mapped to the human GRCh38 transcriptome and total gene transcripts quantified with Salmon (77), significant differential gene expression was calculated using a negative binomial distribution implemented in edgeR (78), Gene Ontology enrichment was performed against terms from the MSigDB (79) while adjusting for inter-gene correlation using the Camera algorithm (80), and significant splice variants were predicted with DRIMseq (81). The significant differentially expressed genes from the ARMOR workflow were then used as input to the signaling pathway impact analysis (SPIA) algorithm to enrich differentially expressed genes against intracellular signaling pathways from five databases including KEGG, Panther, BioCarta, Reactome, and NCI (20,82–85). Differentially expressed genes and splice variants calculated by ARMOR and DRIMSeq were evaluated by the effect measures log2 fold change and likelihood ratio respectively. Confidence in results was accomplished using false discovery-rate adjusted p-values.

### Additional analysis and visualization of differentially expressed genes and gene ontologies

The PRISMA flowchart template was used to generate figure one consistent with transparent reporting of meta-analysis generation and results (86).

The R package ggplot was used to construct the Fig 2 volcano plot from using FDRs and log2 fold change values for each gene from the edgeR output (87).

The KEGG ontology was extracted from the Brite Hierarchy using existing code (82). Genes included in the Brite Hierarchy were then computationally matched to their corresponding edgeR log_2_ fold change values. A statistical enrichment of the KEGG gene ontologies was performed using the R package bc3net (88) prior to visualizing the bc3net enrichment results with the R package Treemap in Fig 3 (89).

### Biomarker prediction using differentially expressed gene data

Salmon-derived transcript counts were organized into a tabular format and samples were randomly assigned to either the testing set (30%) or the training set (70%). The R package randomForest was used to run a supervised classification analysis to determine biomarkers (90). The initial results from the whole transcriptome were then reduced to the 3, 5, and 10 best-scoring transcriptional biomarkers, based on the mean Gini impurity decrease values for each of the features. These values were then sorted by size to determine the transcribed genes from the original dataset with the largest association. The area under the curve (AUC) was calculated from the receiver operator characteristic curves that were generated for each set of random forest results to determine the efficacy of the selected biomarkers for disease prediction.

### Drug prediction using differentially modulated pathways

Drug prediction was conducted by running the significantly modulated pathways file that was generated by SPIA file through Pathways2Targets2.R algorithm (91). To summarize drug findings, the Pathways2Targets output was processed using a custom R script most_common_treatments_2021_09_19.R (92).

## Other information

This meta-analysis was not registered. The review protocol was not prepared separately but is described in detail in the methods section. This meta-analysis received no specific financial support and was supported by general funding from Brigham Young University. Brigham Young University played no role in the ideation, synthesis, or analysis of this transcriptomic meta-analysis. The authors each declare that they have no competing interests. All raw data analyzed in this study can be found in NCBI’s GEO. Analytical codes can be found as cited in materials and methods. All processed data are available in the supplementary materials or at DOI: 10.5281/zenodo.4757764.

## Supporting information

Supplemental Files (Compressed)

## Data Availability

The study ONLY used publicly available human data that were originally located at the Gene Expression Omnibus database, hosted by NCBI (https://www.ncbi.nlm.nih.gov/geo/).

## Acknowledgments

We thank the high-performance computing resources hosted by the BYU Research Computing Center. We also gratefully acknowledge those who generated, provided, and submitted the original data.

## Supporting information captions

**S0 File. B-Cell Non-Hodgkin’s Lymphomas Transcriptomic Meta-Analysis: Supplementary Materials Description**. This document contains a guide to allow readers to easily navigate the supplementary files.

**S1 File. PRISMA 2020 checklist for transparent meta-analysis reporting**. This document contains the PRISMA guidelines for reporting meta-analyses/systematic reviews and the manuscript location of required information.

**S2 File. Differentially expressed gene results (edgeR output)**. The entire BCNHL differentially expressed gene list produced by edgeR. The file is written in TSV format and can be successfully opened in Excel. Some genes in the original TSV file have more associated data than can fit on one line. The genes in the file are ranked according to FDR, with the smallest FDRs at the top. Please note, the file also contains differential expression results for genes which were not significantly differentially expressed at the bottom. For description of column contents, please see S0 File.

**S3 File. Differentially expressed splice variants (by gene; DRIMSeq output)**. The BCNHL differentially expressed splice variant results (by gene) produced by DRIMSeq. Genes are ranked according to adjusted p-value, with the lowest adjusted p-value at the top. NOTE: Should you desire to discover which transcripts of a certain gene are present in the BCNHL dataset, see the “tx_ids” column in Supplementary File S1. For description of column contents, please see S0 File.

**S4 File. Differentially expressed gene ontology results (Camera output)**. The BCNHL differentially expressed gene ontology results produced by Camera. No gene ontologies were significant after performing the FDR correction. For description of column contents, please see S0 File.

**S5 File. Differentially regulated pathway results (SPIA output)**. The BCNHL differentially expressed pathway results produced by SPIA. For description of column contents, please see S0 File.

**S6 File. Drug prediction results by gene (Pathways2Targets unsorted output)**. The raw drug prediction results from Pathways2Targets2.R.

**S7 File. Drug prediction results sorted by most significant pathways impacted (Pathways2Targets sorted output)**. The sorted drug prediction results, ranked according to which drugs impact the highest number of significantly modulated pathways.

**S8 File. Biomarker prediction results (randomForest output)**. The ranked random forest biomarker prediction results. Sheet one contains all genes, and sheet two contains the random forest results when the selection was narrowed to the top three genes.

