## Supplemental Files (Compressed) for "Transcriptomic meta-analysis of non-Hodgkin’s B-cell lymphomas reveals reliance on pathways associated with the extracellular matrix": S0_File.docx

**Supplementary Materials Description**

**S1 File. PRISMA 2020 checklist for transparent meta-analysis reporting.**

File name: S1_File.docx

File description: This document contains the PRISMA guidelines for reporting meta-analyses/systematic reviews and the manuscript location of required information.

**S2 File. Complete differentially expressed gene results (edgeR output).**

File name: S2_File.txt

File description: The entire BCNHL differentially expressed gene list produced by edgeR. The file is written in TSV format and can be successfully opened in Excel. Some genes in the original TSV file have more associated data than can fit on one line.

The genes in the file are ranked according to FDR, with the smallest FDRs at the top.

Please note, the file also contains differential expression results for genes which were not significantly differentially expressed at the bottom.

| **Column Name** | **Column Content Description** |
| --- | --- |
| seqnames | Chromosome upon which this gene is located |
| start | Approximate starting nucleotide on chromosome |
| end | Approximate ending nucleotide on chromosome |
| width | Number of nucleotides in gene |
| strand | Direction gene is read: sense (+) or anti-sense (-) |
| gene_id | Ensembl gene ID |
| gene_name | the HGNC-approved gene symbol; you can search the gene symbol on HGNC's website (https://www.genenames.org/) for more information. |
| gene_biotype | Tells whether gene is protein coding, a pseudo-gene, etc. |
| seq_coord_system | This should say “chromosome” all the way down |
| description | brief description including full gene name |
| gene_id_version | Gene ID version |
| symbol | the HGNC-approved gene symbol; you can search the gene symbol on HGNC's website (https://www.genenames.org/) for more information. |
| entrezid | The gene’s entrez ID |
| tx_ids | List of all splice variants detected in dataset |
| logFC | Log-base-2-fold change; positive means upregulated in BCNHL as compared to healthy, negative means downregulated in BCNHL as compared to healthy. |
| logCPM | logCPM |
| F | F |
| PValue | The p-value is the probability that the result is a false-positive. P-values below 0.05 are typically considered to be significant. |
| FDR | This is the p-value after performing the false-discovery rate (FDR) correction (gives you a more stringent p-value, lower likelihood of false positives). |
| mlog10PValue | The mlog10PValue |

**Supplementary File S3.** **Differentially expressed splice variants (by gene; DRIMSeq output).**

File name: S3_File.xlsx

File description: The BCNHL differentially expressed splice variant results (by gene) produced by DRIMSeq. Genes are ranked according to adjusted p-value, with the lowest adjusted p-value at the top.

NOTE: Should you desire to discover which transcripts of a certain gene are present in the BCNHL dataset, see the "tx_ids" column in Supplementary File S1.

| **Column Name** | **Column Content Description** |
| --- | --- |
| gene_id | Ensembl gene ID |
| description | brief description including full gene name |
| symbol | the HGNC-approved gene symbol; you can search the gene symbol on HGNC's website (https://www.genenames.org/) for more information. |
| lr | likelihood ratio |
| df | # of *alternate* transcripts present; df + 1 = total # of splice variants present in the dataset |
| pvalue | The p-value is the probability that the result is a false-positive. P-values below 0.05 are typically considered to be significant. |
| adj_pvalue | This is the p-value after performing the Benjamini-Hochberg correction for multiple testing (gives you a more stringent p-value, lower likelihood of false positives). |
| mlog10PValue | The mlog10PValue |

**Supplementary File S4.** **Differentially expressed gene ontology results (Camera output).**

File name: S4_File.xlsx

File description: The BCNHL differentially expressed gene ontology results produced by Camera. No gene ontologies were significant after performing the FDR correction.

| **Column Name** | **Column Content Description** |
| --- | --- |
| GeneSet | The name of the gene set analyzed in this row. To learn more about a particular gene set, search it on the MSigDB website (https://www.gsea-msigdb.org/gsea/msigdb/). |
| NGenes | Total number of genes included in this gene set. |
| Correlation | A sliding scale telling how closely your data set correlates with this gene set. 0 indicates that there is no correlation (in other words, the relationship is random), and +1 or -1 indicates that the correlation is perfect (no randomness in relationship). |
| Direction | Whether the gene set was upregulated or downregulated. |
| PValue | The p-value is the probability that the result is a false-positive. P-values below 0.05 are typically considered to be significant. |
| FDR | This is the p-value after performing the false-discovery rate (FDR) correction (gives you a more stringent p-value, lower likelihood of false positives). |

**Supplementary File S5.** **Differentially regulated pathway results (SPIA output).**

File name: S5_File.xlsx

File description: The BCNHL differentially expressed pathway results produced by SPIA.

| **Column Name** | **Column Content Description** |
| --- | --- |
| Rank | Results are ranked by pGFWER (the most stringently adjusted p-value) |
| Name | Pathway name |
| pSize | Number of genes in pathway |
| NDE | Number of pathway genes that are differentailly expressed in this dataset |
| pNDE | How significant the pSize:NDE ratio is |
| tA | measure of change between healthy and lymphoma expression; directionality indicates up- or down-regulation |
| pPERT | An adjusted P-value |
| pG | An adjusted P-value |
| pGFdr | An adjusted P-value |
| pGFWER | The most stringently-adjusted p-value |
| Status | Tells whether pathway is upregulated or downregulated |
| SourceDB | The database from which this pathway originated |

**Supplementary File S6. Drug prediction results by gene (Pathways2Targets unsorted output).**

File name: S6_File.xlsx

File description: The raw drug prediction results from Pathways2Targets2.R.

| **Column Name** | **Column Content Description** |
| --- | --- |
| Target_ID | ENSEMBL gene ID of the targeted gene. |
| Target_Symbol | HGNC gene symbol. |
| Target_Name | Expanded gene name. |
| Drug_ID | CHEMBL drug ID. |
| Drug_Name | Name of drug. |
| Is_FDA_Approved | Tells whether this drug is FDA-approved. |
| Highest_Clinical_Trial_Phase | Tells clinical trial phase. Level 4 is the best. |
| Has_Been_Withdrawn | Tells whether drug was withdrawn. Avoid withdrawn drugs for most applications. |
| Pathway_DB | Which database the significantly modulated pathway containing this gene came from. |
| Pathway_Name | The name of the significantly modulated pathway containing this gene. |

**Supplementary File S7.** **Drug prediction results sorted by most significant pathways impacted (Pathways2Targets sorted output).**

File name: S7_File.xlsx

File description: The sorted drug prediction results, ranked according to which drugs impact the highest number of significantly modulated pathways.

| **Column Name** | **Column Content Description** |
| --- | --- |
| Drug_ID | CHEMBL drug ID. |
| Drug_Name | Name of drug. |
| Significant_Pathways_Targeted | Tells how many significantly modulated pathways this drug targets through known mechanisms. |
| Is_FDA_Approved | Tells whether this drug is FDA-approved. |
| Highest_Clinical_Trial_Phase | Tells clinical trial phase. Level 4 is the best. |
| Has_Been_Withdrawn | Tells whether drug was withdrawn. Avoid withdrawn drugs for most applications. |

**Supplementary File S8.** **Biomarker prediction results (randomForest output).**

File name: S8_File.xlsx

File description: The ranked random forest biomarker prediction results. Sheet one contains all genes, and sheet two contains the random forest results when the selection was narrowed to the top three genes.

| **Column Name** | **Column Content Description** |
| --- | --- |
| Features | The name of the gene tested by randomForest. |
| Mean_Decrease_Gini | The ranking criteria of randomForest results. Bigger is better. |
